# Introduction of the topic of snake bite and envenomation in medical curriculum using a problem-based learning approach

**DOI:** 10.1101/2024.09.18.24313523

**Authors:** Anita Devi Krishnan Thantry, Wai Marr Lin, Raja Norazura Raja Zahadi

## Abstract

**Introduction:** Snake bite is a neglected tropical disease of Asia and African countries. The risk factors, disease burden, pathology, clinical management aspects of the condition deserve to be emphasized in the medical curriculum especially in the tropical countries. In Malaysia, there are at least 18 different species of venomous land snakes among which cobra bites and Malayan pit viper bites predominate. The coverage of snake bite envenomation in medical school curriculum is inadequate judging from the poor knowledge among health care providers. PBL is an instructional mode where “triggers” from a problem case is used to define learning objectives. PBL enables students to understand the relevance of underlying scientific knowledge and principles in clinical practice.

**Methods:** We attempted the delivery of topic of snake bite to 100 students in Year 1 module “Body reactions to Various Agents” using a problem-based learning (PBL) approach. The PBL case was designed; conducted according to standard format. The students were evaluated during the first and second session according to standard format. A questionnaire on the various aspects of the case, group work and facilitator was circulated and survey was based on the Likert scale. Problem based question on the same topic was used in end of block examination to assess knowledge assimilation.

**Results and conclusion:** The study showed a positive response in the students’ attitude, perception and gain of knowledge on the topic of snake bite envenomation. Problem based learning was considered an enjoyable and well-suited model for the delivery of the topic. The paucity of student friendly and medically relevant literature in textbooks and E-resources was highlighted by the students. The student score for the PBL sessions was high while the marks scored for the problem –based question showed a significant positive association between the top-scorers of the year group and the PBQ marks (p > 0.05). The PBL method is proven to improve the affective skills in the students while the cognitive skills improvement is not enhanced.

## Introduction

Venomous snakes are found throughout the world, except some island nations, frozen environment and high altitudes. The morbidity and mortality due to envenomation in the rural areas is high due to poor access and sub-optimal health services in these regions. The psychological and physical sequale (e.g.: tissue necrosis) in survivors are grave as most affected victims are young and the economic impact is considerable^1^. This neglected tropical disease (NTD) has large health implications in a country like Malaysia which has varied flora and fauna with many common and rare species of snakes being reported^2^.

In Malaysia, there are at least 18 different species of venomous land snakes among which cases of cobra bites and Malayan pit viper bites predominate ^3^. The incidence of death due to snakebite in Malaysia is estimated as 102 per million in year 2024 and it is estimated that between 15K and 55K die from snake bites across the countries in Asia every year^4^.

The risk factors, disease burden, pathology, clinical management aspects of the condition deserve to be emphasized in the medical curriculum especially in the tropical countries. The coverage of the topic of snake bite and envenomation in medical school curriculum is inadequate judging from the poor knowledge among health care providers ^(5,6).^ Training medical personnel on modern management and selective use of anti-venom in snake bite is of great importance in public health and emergency medicine globally^7,8^.

Many medical schools do not have substantive training modules on snakebite included in their curriculums^9^. A student-centered problem-based learning (PBL) approach to the topic would facilitate active learning, which, in addition to gaining medical knowledge, will foster the competencies of practice-based learning and improvement, interpersonal and communication skills, professionalism, and systems-based practice^10^. This study was based on a problem-based module on snake bite where their knowledge, soft skills and perception was assessed after its implementation.

## Materials and Methods

Using the PBL methodology, the topic of snake bite was introduced to first-year students in the block on ‘Infectious agents’. A case of viper bite was constructed which composed of 3 triggers and the learning objectives appropriate for the year group were identified and endorsed in the faculty.

A total of 100 students took part in the study which included 2 sessions each of 2 hours conducted in two consecutive weeks. The students were divided into groups of 7-8 and the sessions were conducted simultaneously by facilitators in ear-marked PBL rooms. During session 1, the students were provided with the triggers following which brainstorming and formulation of learning issues were done. After a week, they re-grouped for session 2 with newly acquired information and discussed their findings and its relevance to the given case. Students were assessed by the facilitator during both sessions based on their contribution using a standardized evaluation form*. The students" consolidated report was assessed by the facilitator and the marks were added to the total score obtained from session 1 and 2. The total PBL marks for the PBL session was given out of a maximum of 40 marks.

A questionnaire survey on their perception towards the case, the facilitator and the availability of resources was assessed at the end of the second session.

The topic knowledge was further evaluated in the end of block exam in which a problem-based question (PBQ) on snake envenomation was incorporated. The question assessed the student’s knowledge on basic first –aid measures to be advocated following snake bite and the various types of viper toxins and their effects on the tissue for a total of 30 marks.

The comparison of the student performance score in PBL as against the total score in the PBQ was attempted.

## Results and Discussion

A total of 100 students from the Year 1, block 3 cohort were enrolled in the study of which 48 were medical students while 52 were dental students (Figure 1).

**Figure 1:**
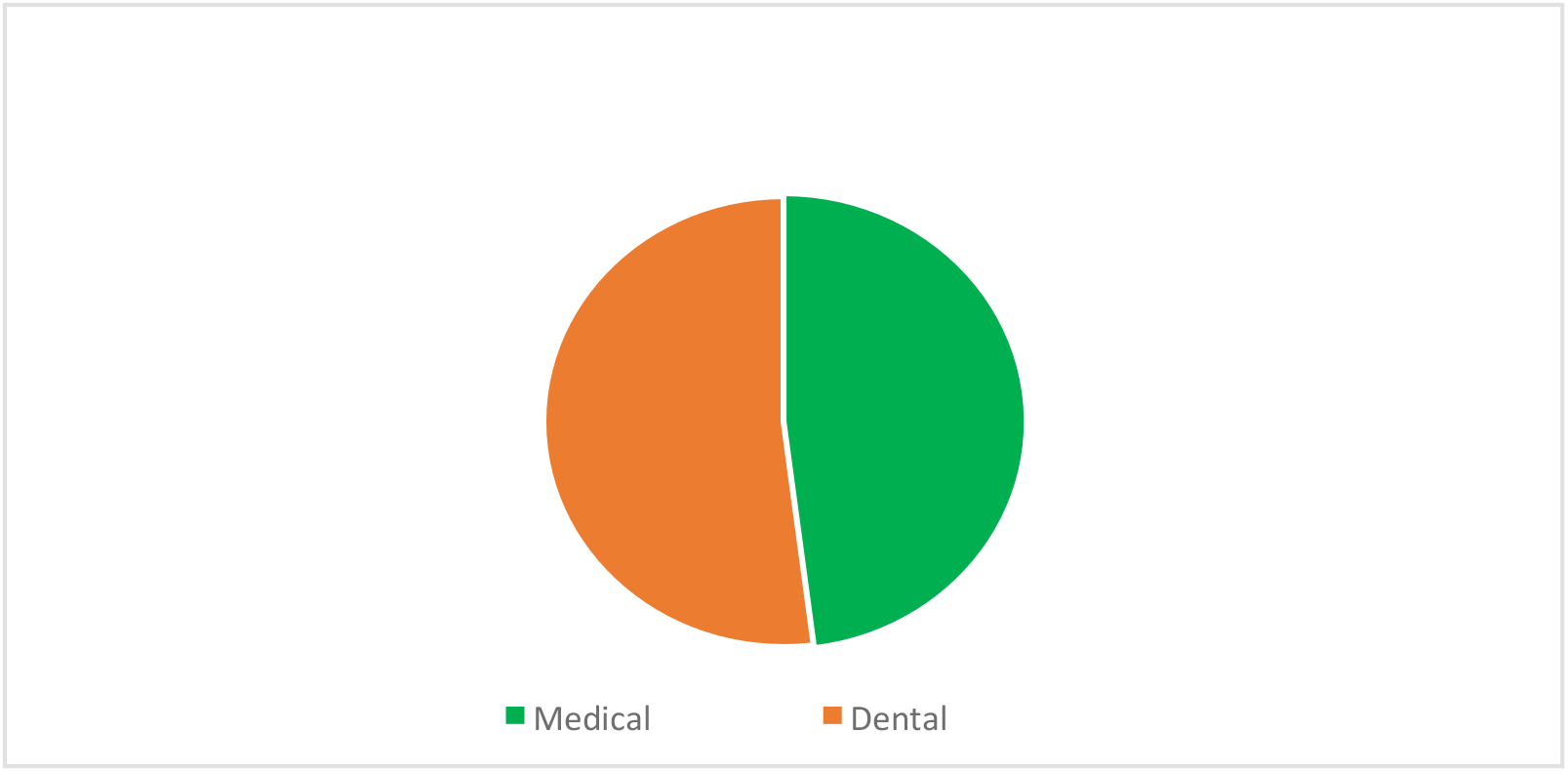
Year 1; Block 3 students participating in the study

The average score for session 1 was 3.5/ 4 while the average score for session 2 was 13/ 14. The final PBL report score on an average was 11/ 12 with the total score for the PBL session being 27.5/30. (Table 1).

**Table 1:** Scores achieved by the students in each session of PBL and for their final report (n=100).

| **PBL on Snake Bite** | **Score** | **Maximum marks** |
| --- | --- | --- |
| Average score for PBL Session 1 | 3.5 | 4 |
| Average score for PBL Session 2 | 13.0 | 14 |
| Average score for PBL report | 11.0 | 12 |
| **Average total score for PBL on Snake bite** | **27.5** | **30** |

The mean score for the PBL session was 91.7% while the average score for the PBQ was 17.4/ 30(58.0%). The average mean score for the final end of block exam was 62.6 % (Table 2).

**Table 2:** Mean scores of the tests assessing the topic of snake bite.

| **Topic of Snake Bite** |  |  |
| --- | --- | --- |
| **Assessment methods (Mean score)** | **Score** | **%** |
| PBL sessions (Total) | 27.5/30 | 91.7 |
| Problem based question (PBQ) | 17.4/30 | 58.0 |
| 'End of block' Exam marks | 62.6/100 | 62.6 |

Thirty-nine students among the total 100 failed in the PBQ (score less than 15/30). There was a positive corelation between the total marks in the end of block exam with the PBQ score (Figure 2) with a p value= 0.57. This shows that the students with a high block exam score also scored high in the PBQ component.

**Figure 2:**
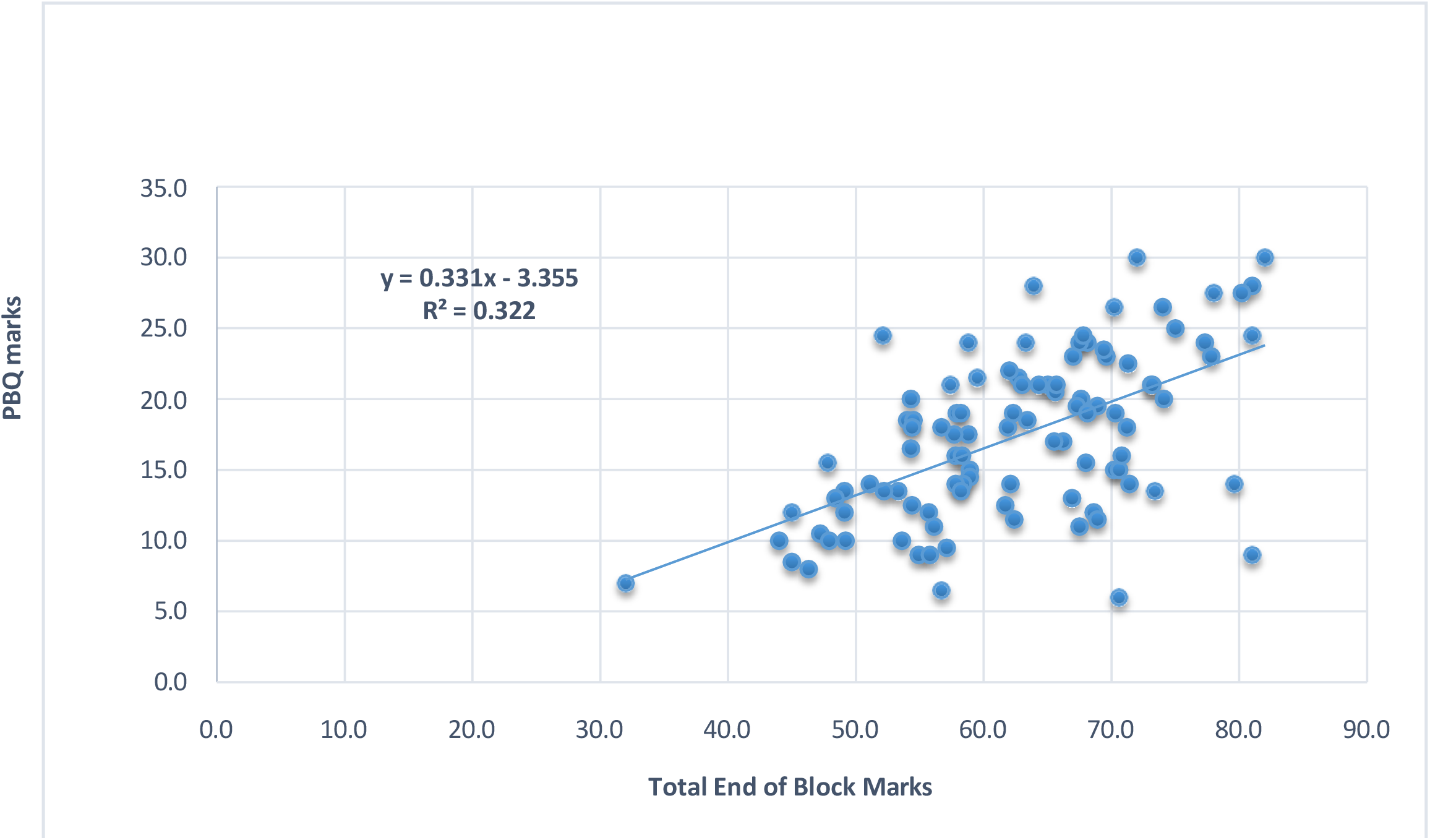
Co-relation of PBQ on snake bite marks with Total Score in the Block Exam

Further the student feedback to the PBL case was evaluated under the following subtitles: PBL case, role of facilitator, availability of resources and the venue for conducting PBL. The case was considered as well suited for the problem-based learning method by most students. (Figure 3)

**Figure 3:**
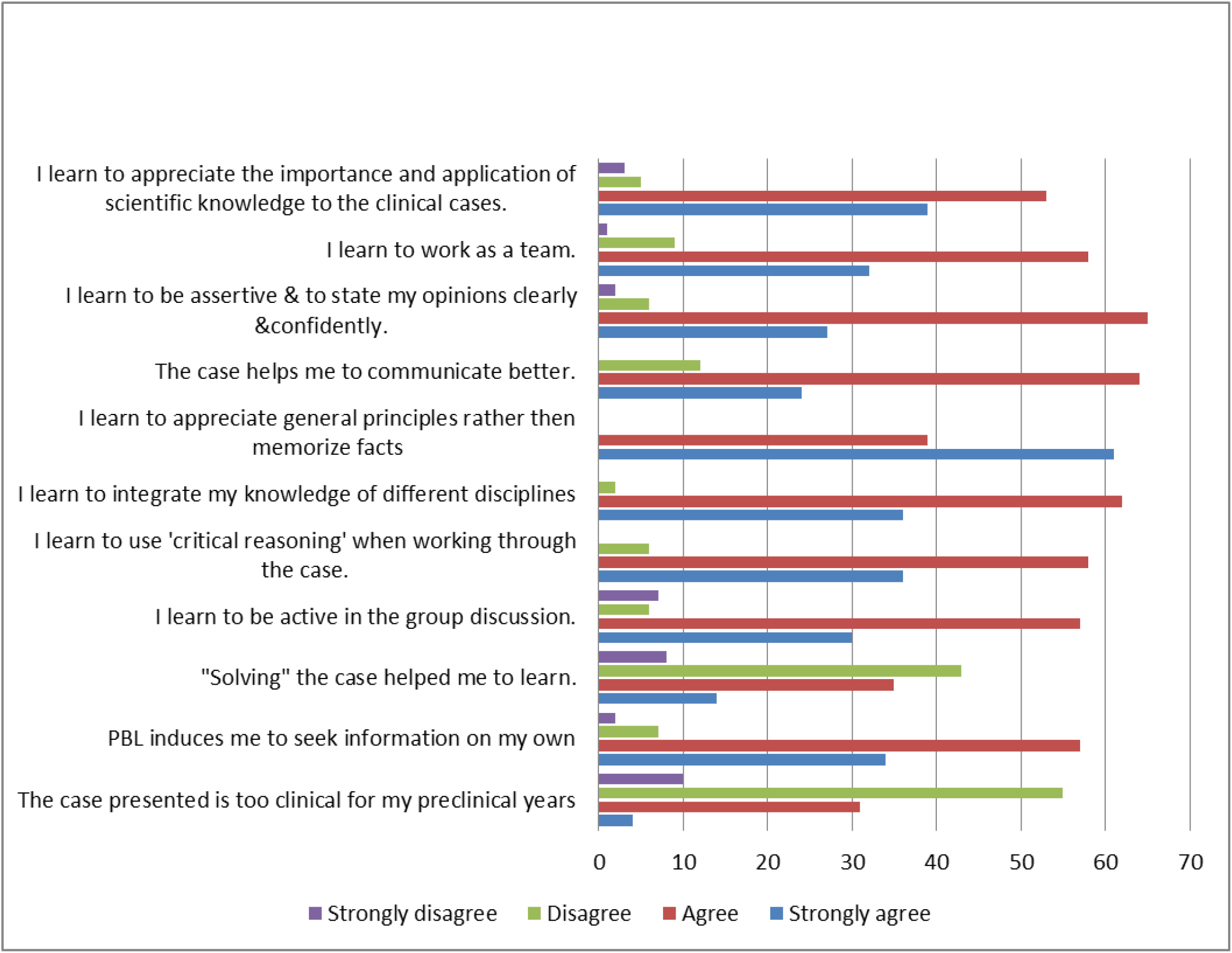
Student perception about PBL Case on Snake Bite

The feedback on the role of facilitator was also positive (Figure 4) and was adequately following the principles of problem-based learning.

**Figure 4:**
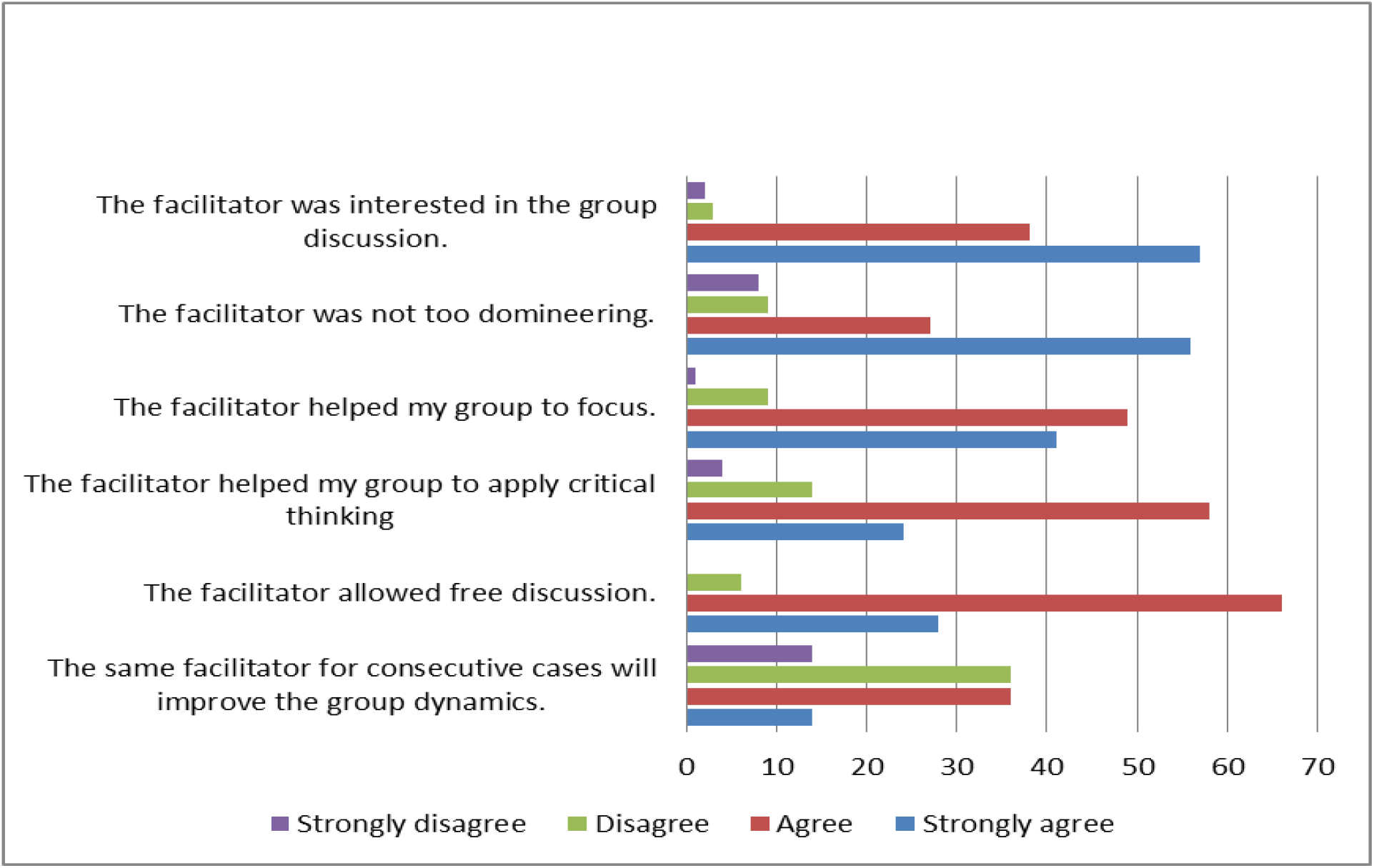
Student perception about facilitators for PBL case on snake Bite

The feedback on resources available for self-directed learning was not very encouraging (Figure 5). The majority felt that availability of books or E materials which focus on the mechanism by which the enzymes and toxins of snake produce their effect on the human tissue as well the principles of anti-venom was inadequate.

**Figure 5:**
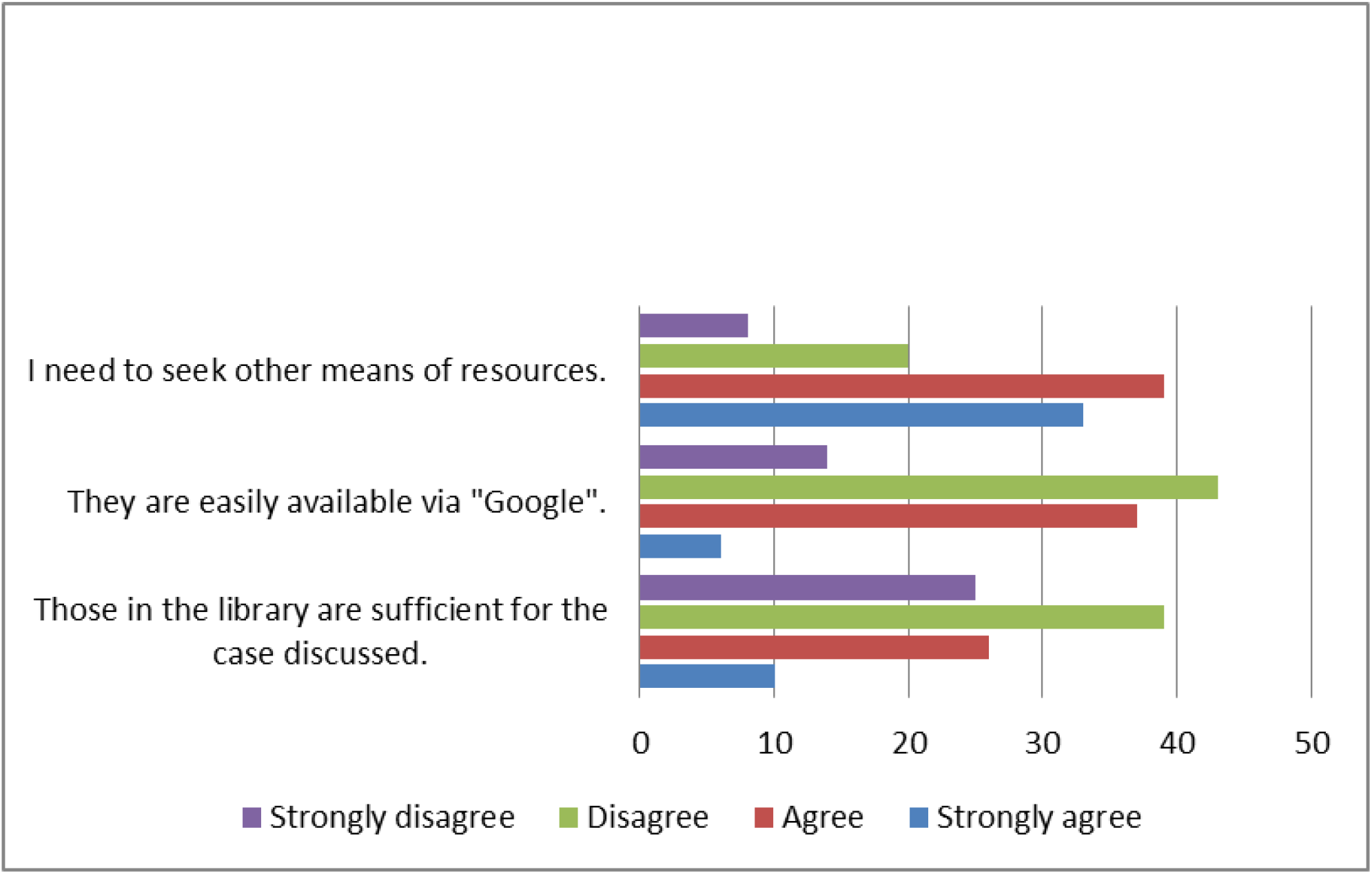
Student perception about availabilty of resources for PBL case on Snake Bite

The facilitators and student comments also reflected the above findings. Some of the blinded facilitator comments included, *“Interesting case for student, triggered researching on different types of snakes in SE Asia “; “Student use online educational videos regarding snake bite”*. Student-blinded response highlighted include-*“Resources not easily available on google”; “Good case which triggered us to read about snake bite and its consequences”; “Case was interesting and enjoyable”*.

## Discussion

The study was conducted on 100 students of Year 1, block 3 which included 48 medical and 52 dental students. The topic of snake bite when introduced in the problem-based format, has generated a keen interest by encouraging the students to search for information and share the resource among their peers. In addition to gaining knowledge, they had the opportunity to develop soft skills of communication and team effort. The study also proves that even though the students had the opportunity to learn and assimilate their knowledge over two sessions of PBL (total of 4hours in addition to self-directed learning) it did not reflect as a better score among students who had overall low grades in the final examination. This supports the fact that PBL helps more to foster the affective (soft) skills while the same topic when tested in the exam, does not reflect on the cognitive skill^11^.

In addition, the topic of snake bite and envenomation was considered a difficult subject to research on which is reflected in the feedback on the scarce availability of resources (both in the library books as well as on the internet)^12^. This points to an urgent need to provide better resources on this neglected tropical disease, which may be due to higher incidence of this condition in the tropics. The World Health Organization (WHO) recommends that education and training in the prevention and management of snakebites should be included in the curriculum of medical and nursing schools^13^. As snakebite is a clinical problem mainly presented in primary care, medical schools should consider revising their curriculum to include snakebite management during Emergency Medicine and Family Medicine clinical postings rather than an hour of didactic lecture in preclinical phase ^14^.

## Conclusion

Problem based learning method is a widely used learning strategy for delivery of subjects which are not well – covered in the curriculum. It fosters soft skills of improving interpersonal communication, teamwork, analytical reasoning and self –directed learning which are essential traits required to be developed in a medical student. Snake bite and envenomation requires to be introduced to medical students in pre-clinical years to focus on the hematological and neurological pathology followed by revisit in the clinical years. More text-based and student friendly peer-reviewed information should be made available in medical books and other commonly accessed sources of medical information.

## Data Availability

All relevant data are within the manuscript and its Supporting Information files.

## References

1. Bittenbinder, M.A., van Thiel, J., Cardoso, F.C. et al.; 2024. Tissue damaging toxins in snake venoms: mechanisms of action, pathophysiology and treatment strategies. Commun Biol 7, 358 (2024). 10.1038/s42003-024-06019-6

2. WHO. Control of Neglected Tropical Diseases 2012; https://www.who.int/teams/control-of-neglected-tropical-diseases/snakebite-envenoming/treatment

3. Ismail, A.K., Wah, T.E., Das, I., Vasaruchapong, T. and Weinstein, S.A., 2017. Land snakes of medical significance in Malaysia. Malaysia Biodiversity Information System (MyBIS).

4. World Population Review. Snake Bite Deaths by Country 2024. https://worldpopulationreview.com/country-rankings/snake-bite-deaths-by-country

5. Afroz A, Siddiquea BN, Shetty AN, Jackson TNW, Watt AD. Assessing knowledge and awareness regarding snakebite and management of snakebite envenoming in healthcare workers and the general population: A systematic review and meta-analysis. PLoS Negl Trop Dis. 2023 Feb 9;17(2): e0011048. doi: 10.1371/journal.pntd.0011048. PMID: 36757933; PMCID: PMC9910687.

6. Godpower Chinedu Michael, Auwal Adam Bala, Mustapha Mohammed. Snakebite knowledge assessment and training of healthcare professionals in Asia, Africa, and the Middle East: A review, Toxicon: X, Volume 16, 2022

7. Sapkota S, Pandey DP, Dhakal GP, Gurung DB (2020) Knowledge of health workers on snakes and snakebite management and treatment seeking behavior of snakebite victims in Bhutan. PLoS Negl Trop Dis 14(11): e0008793. 10.1371/journal.pntd.0008793

8. Eleanor Strand, Felipe Murta, Anna Tupetz, Loren Barcenas, Ashley J. Phillips, Altair Seabra Farias, Alícia Cacau Santos, Gisele dos Santos Rocha, Catherine A. Staton, Flávia Regina Ramos, Vinícius Azevedo Machado, Fan Hui Wen, João R.N. Vissoci, Jacqueline Sachett, Wuelton Monteiro, Charles J. Gerardo. Perspectives on snakebite envenoming care needs across different sociocultural contexts and health systems: A comparative qualitative analysis among US and Brazilian health providers, Toxicon: X, Volume 17, 2023, 100143

9. Bhaumik S, Zwi AB, Norton R, Jagnoor J. How and why snakebite became a global health priority: a policy analysis. BMJ Glob Health. 2023 Aug;8(8): e011923. doi: 10.1136/bmjgh-2023-011923. PMID: 37604596; PMCID: PMC10445399.

10. Lim WK. Problem Based Learning in Medical Education: Handling Objections and Sustainable Implementation. Adv Med Educ Pract. 2023 Dec 28; 14:1453–1460. doi: 10.2147/AMEP.S444566. PMID: 38164409; PMCID: PMC10758192.

11. Ribeiro, Luis Roberto C., The Pros and Cons of Problem-Based Learning from the Teacher’s Standpoint, Journal of University Teaching & Learning Practice, 8(1), 2011. Available at: http://ro.uow.edu.au/jutlp/vol8/iss1/4

12. Bonilla-Aldana DK, Bonilla-Aldana JL, Ulloque-Badaracco JR, et al. Snakebite-Associated Infections: A Systematic Review and Meta-Analysis. The American Journal of Tropical Medicine and Hygiene. 2024;110(5):874–886. doi:10.4269/ajtmh.23-0278

13. Qiu, C., Qiu, XF., Liu, JJ. et al. An effective snakebite first aid training method for medics in the Chinese troops: a RCT. Military Med Res 6, 39 (2019). 10.1186/s40779-019-0230-9

14. Pandey DP, Khanal BP. Inclusion of incorrect information on snake bite first aid in school and university teaching materials in Nepal. J of Toxicology and Environmental Health Sciences;2013, 5(3):43–51

